# Report-Guided Semi-Supervised Learning for Scalable Prostate Cancer Detection on Biparametric MRI: Multicenter Prospective Validation and Multimodal Integration

**DOI:** 10.64898/2026.08.05.26359781

**Authors:** A. Calado, J. G. de Almeida, A. S. C. Verde, M. Tsiknakis, K. Marias, D. Regge, N. Papanikolaou, the ProCAncer-I Consortium

## Abstract

**Purpose:** To prospectively validate a semi-supervised learning framework with a lesion-only teacher model (RG-SSL-LOC) for scalable clinically significant prostate cancer detection on biparametric MRI (bpMRI) and assess its added value in multimodal models.

**Materials and Methods:** A multicenter dataset of 13,706 bpMRI examinations (13,630 patients, 27 centers) was used for model development/validation. Three segmentation models (fully supervised learning [FSL], a state-of-the-art report-guided semi-supervised approach [RG-SSL], and the proposed RG-SSL-LOC) were evaluated at lesion- and case-level on external retrospective, external prospective, and internal prospective cohorts. Predictions from the best-performing model were combined with clinico-radiologic variables in a multimodal approach. All case-level results were compared with PI-RADS.

**Results:** At lesion level, RG-SSL-LOC achieved higher median Dice than FSL and RG-SSL (0.49 vs 0.41 and 0.40; both *p*<.001). At case level, RG-SSL-LOC achieved area-under-the-curve (AUC) values of 0.83, 0.82, and 0.87 in the external retrospective, external prospective, and internal prospective cohorts, respectively. Compared with FSL, AUCs were 0.84 (*p*=.237), 0.80 (*p*=.020), and 0.84 (*p*<.001); compared with RG-SSL, AUCs were 0.83 (*p*=.929), 0.82 (*p*=.652), and 0.86 (*p*=.007); compared with PI-RADS, AUCs were 0.78 (*p*=.055), 0.83 (*p*=.652) and 0.86 (*p*=.480). Combined with clinico-radiological variables, RG-SSL-LOC significantly improved AUC versus clinico-radiological variables alone in the external retrospective (0.85 vs 0.80, *p*=.002), external prospective (0.87 vs 0.84, *p*=.008), and internal prospective (0.91 vs 0.88, *p*<.001) cohorts; in the latter, it reduced unnecessary biopsies by 15.19%.

**Conclusion:** RG-SSL-LOC achieves better segmentation quality than other methods, demonstrates robust prospective multicenter performance and improves multimodal detection.

**Summary:** A report-guided semi-supervised method outperforms fully-supervised baseline on prospective multicenter data for prostate cancer detection and adds value in multimodal approaches, effectively using unlabelled data and facilitating model scaling.

**Key Points:**

- The use of a lesion-only teacher model in a state-of-the-art report-guided semi-supervised learning framework improved prostate cancer segmentation quality and detection performance in biparametric MRI
- Report-guided semi-supervised learning more than tripled the amount of training data available, with the proposed lesion-only teacher approach retaining 7.02% more malignant cases than the state-of-the-art approach.
- In an internal prospective cohort, the proposed multimodal approach could potentially reduce unnecessary biopsies by 15.2%.

## Introduction

Pseudolabelling is a commonly used semi-supervised (SSL) method based on using a *teacher* model trained on labeled data to generate pseudolabels for unlabeled instances, increasing the amount of data available for training a *student* model. Previous studies on prostate cancer (PCa) detection have refined naïve pseudolabelling by incorporating additional information such as lesion count or location. Bosma *et al*. (2023) (1) used automatic radiology report processing to incorporate the number of Prostate Imaging Reporting and Data System (PI-RADS) > 3 lesions: this was used to filter the teacher model output such that the number of inferred lesions was identical to the number of lesions with PI-RADS>3, leading to a student model which outperformed a fully supervised (FSL) baseline by 2% case-level area-under-the-curve (AUC). Building on this work, Chen *et al*. (2024) (2) incorporated location information extracted from radiology reports, showing that, when compared with FSL, location-based SSL led to comparable lesion segmentation performance with a reduced false positive rate. Finally, Bosma *et al*. (2025) (3) further improved this by automating and refining lesion location extraction with Scalable Clinical Annotation with Location Evidence (SCALE). Using radiology and pathology reports, SCALE automatically extracts lesion locations using large language models. When compared with the PI-CAI-1 AI system (the ensemble of the top 5 submissions to the PI-CAI challenge (4) trained on 10,207 cases), SCALE improved its clinically significant prostate cancer (csPCa) detection by 0.006 case-level AUC, leveraging data from more than 16,000 cases. While significant in their developments, these methods have significant limitations. First, performance was not evaluated prospectively, raising questions about long-term performance. Second, multimodal incorporation with clinical data was not extensively evaluated. Third, PCa detection was not considered. The latter is particularly relevant: while patients might not be flagged for csPCa, they still require active surveillance to ensure no disease progression.

To address these limitations, we build upon SCALE by introducing four key contributions: 1) prospective multicenter validation; 2) multimodal integration with clinico-radiological variables; 3) csPCa vs. PCa comparison; 4) a lesion-only teacher model trained exclusively on ISUP (International Society of Urological Pathology) ≥ 1 cases to generate more accurate location-corrected pseudolabels.

## Materials and Methods

### Dataset

The models reported in this study were developed/validated on a large multicentric bpMRI dataset comprising T2-weighted (T2w), apparent diffusion coefficient (ADC), and diffusion-weighted imaging (DWI) sequences. The full dataset includes 13,706 cases (13,630 patients) from 27 centers in 10 countries and 2 continents and is composed of three public — PI-CAI (4) (1,500 cases, 1,476 patients), PROMIS (5) (575 cases, 575 patients) and UCLA (6) (837 cases, 837 patients) — and one private dataset — ProstateNET (7) (10,794 cases, 10,742 patients). The latter includes 8,188 cases (8,167 patients) collected retrospectively and 2,606 (2,575 patients) prospectively, before and after March 31 2022, respectively. Since all cases from PI-CAI, PROMIS and UCLA were collected prior to this date, they were treated as retrospective data for the context of this study. The inclusion criteria were: 1) age ≥18 at the time of diagnosis; 2) bpMRI including T2w imaging, high *b* value DWI and ADC; 3) Biopsy/prostatectomy histology results or follow-up of ≥ 1 year for negative cases.

All negative cases in the UCLA and PROMIS datasets were confirmed histologically, whereas negative cases in ProstateNET and PI-CAI were confirmed either histologically or through follow-up (≥ 1 year and ≥ 3 years, respectively) with no evidence of disease. PCa and csPCa were defined as ISUP ≥ 1 and ≥ 2, respectively. Lesion location information was available for PROMIS and ProstateNET, described using Barzell zones and the PI-RADS v2 sector map, respectively. UCLA included biopsy coordinates, each associated with an ISUP grade. PI-RADS scores were only available for ProstateNET. Manual lesion segmentations were available for PI-CAI (csPCa only) and UCLA, as well as for a subset of ProstateNET retrospective data (816 cases, 815 patients, 8 centers), PNET-INT-RETRO-SEG hereafter.

### Data Splitting

The dataset was divided according to Figure S1. Labeled training data (*i*.*e*. with manual lesion segmentations) consisted of UCLA, PNET-INT-RETRO-SEG, and a subset of PI-CAI (PI-CAI-TRAIN) consisting of cases from Radboud University Medical Center and Ziekenhuis Groep Twente. Unlabeled training data included PROMIS and the remaining retrospective ProstateNET cases (6,845 cases from 6,831 patients) — PNET-INT-RETRO hereafter — from the same eight centers included in PNET-INT-RETRO-SEG

The remainder of the data was reserved for lesion- and case-level validation. Lesion-level validation was performed on Prostaat Centrum Noord-Nederland cases (338 patients) from PI-CAI (PI-CAI-VAL), for which only csPCa lesion segmentations were available. For this reason, lesion-level evaluation was restricted to csPCa and PI-CAI cases were excluded from the PCa detection task.

Case-level validation was conducted on the remaining ProstateNET dataset, which was divided into three cohorts: 1) PNET-INT-PROS, an internal prospective cohort of 1,954 cases (1,926 patients) from eight of the centers used for model training; 2) PNET-EXT-RETRO, an external retrospective cohort of 527 cases (527 patients) from four centers not included in training; 3) PNET-EXT-PROS, an external prospective cohort of 652 cases (649 patients) from five centers not included in training — four of which included in PNET-EXT-RETRO. Data preprocessing is described in the supplementary materials.

### Experiments

We trained three csPCa segmentation models — one FSL and two SSL — using a cross-validation (CV) strategy (Figure S2 [a]). For PCa segmentation, only the FSL model and the best-performing SSL approach were considered. Both SSL models used the same teacher-student framework as SCALE, in which a teacher model trained on labeled data is used to generate pseudolabels for unlabeled cases. These pseudolabels are then corrected using report-guided information (Figure 1).

**Figure 1.**
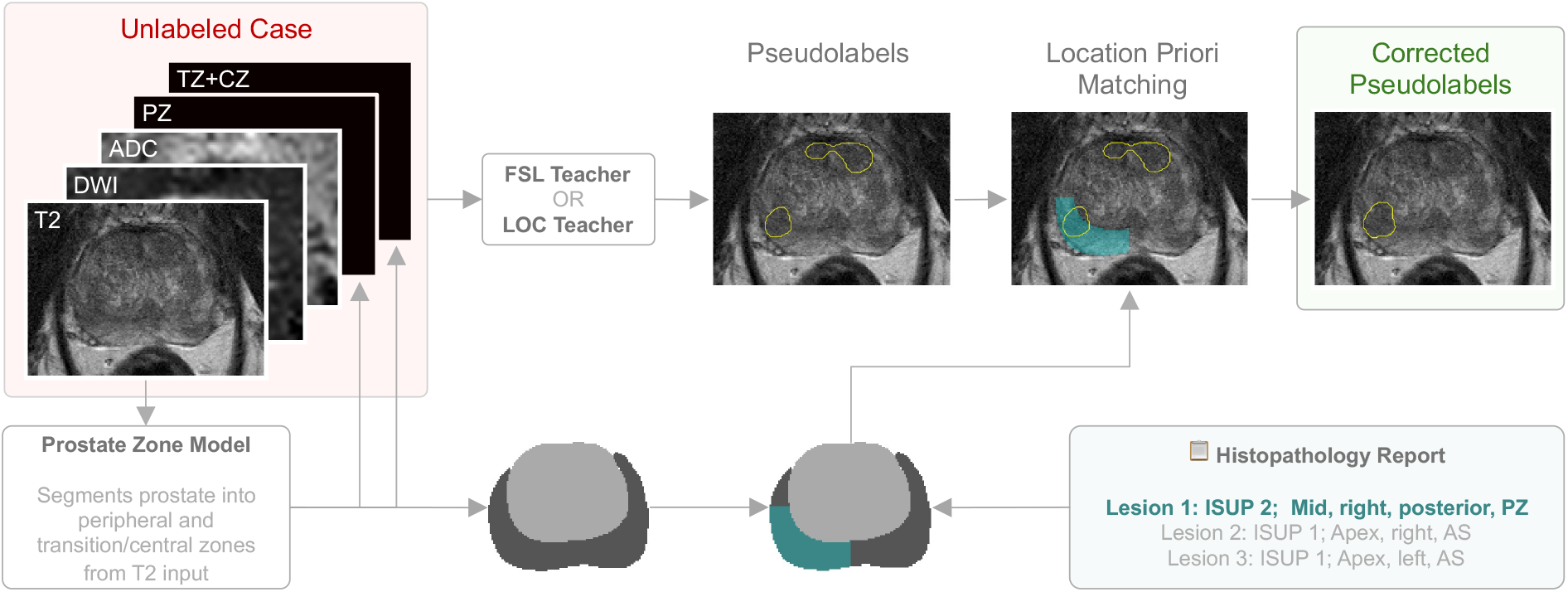
Example of a report-guided correction. First, a prostate zonal segmentation model produces masks of the prostate zones from T2 and adds them as two additional input channels. Second, an unlabeled case is processed by a teacher model to generate pseudolabels (yellow). Third, the zone masks are combined with information from the histopathology report to estimate voxel-level location priors (cyan). Fourth, the pseudolabels are then refined by retaining only those that spatially overlap with location priors, while unmatched pseudolabels are discarded.

First, segmentation masks are generated for the peripheral and central/transition zones using an in-house zone segmentation model (8). Next, for each unlabeled positive case, the teacher model generates pseudolabels. The zonal masks are combined with the report-derived lesion locations, as well as histopathology findings, to generate voxel-level location priors and assign the corresponding ISUP grades. Prostate sectors were estimated using simple heuristics: left/right was defined by the sagittal plane centered in the whole-gland mask, anterior/posterior by the coronal plane centered in the whole-gland mask, apex/mid/base by dividing the prostate into equal thirds along the axial plane and peripheral/transition/central according to the corresponding masks. Each pseudolabel is then matched with a location prior based on highest overlap. Matched pseudolabels are kept and unmatched ones are excluded. If all pseudolabels are excluded (*i*.*e*., zero overlap across all location priors), the case is excluded altogether from training, in accordance with the findings from Bosma *et al*. (2025). Finally, the student model is trained with both labeled and pseudolabeled cases.

The fundamental difference between the two SSL approaches lies on the teacher model. The first approach — RG-SSL hereafter —, uses a csPCa/PCa segmentation FSL model, depending on the task, trained on both positive and negative cases as the teacher (as in Bosma et al. [2025]). In contrast, the proposed method — RG-SSL-LOC (lesion-only correction) —, uses a PCa segmentation FSL model trained exclusively on positive cases as the teacher for both tasks, LOC teacher hereafter.

The multimodal models were based on logistic regression (LR) ensembles incorporating the output of the corresponding best-performing segmentation model with clinico-radiological variables: PSA (Prostate-Specific Antigen) level, age, PI-RADS score, and lesion location (Figure S2 [b]). To evaluate the incremental value of the segmentation-based predictions, additional ensembles were trained using only clinico-radiological variables (*i*.*e*., excluding the segmentation model output). To quantify the relative importance of each covariate, the LR coefficients and their corresponding 95% confidence intervals (CIs) were averaged across ensemble models.

The csPCa segmentation models were evaluated on PI-CAI-VAL at lesion-level using DSC (Dice Similarity Coefficient) and fROC (free-response Receiver Operating Characteristic) analysis. For the fROC analysis, a candidate lesion was considered a true positive if its intersection-over-union with a ground-truth connected component exceeded 0.1. All models were evaluated at case-level on the external retrospective/prospective and internal prospective cohorts using AUC, sensitivity at PI-RADS>3 specificity and specificity at PI-RADS>3 sensitivity. The latter provides a measure of the potential reduction of unnecessary biopsies while maintaining a detection rate comparable to standard radiological assessment.

We conducted a subgroup analysis for the baseline FSL and best-performing SSL segmentation models on the external cohorts to better understand the performance differences between the retrospective and prospective distributions. Performance was evaluated in terms of case-level AUC across subgroups defined by acquisition center, scanner manufacturer (General Electric, Philips, and Siemens), magnetic field strength (1.5T and 3.0T), age, and PSA level. Age and PSA were each divided into three groups using the first and third quartiles of the training data as cut-off points. Finally, a decision curve analysis (DCA) was performed for the best-performing multimodal model.

The 95% CIs were computed with the DeLong’s method and bootstrap resampling (10,000 samples) for AUC and specificity, respectively. Statistically significant differences between AUCs were assessed using DeLong’s test, while differences in sensitivity and specificity values, along with DSC, were evaluated using the Wilcoxon signed-rank test (*alpha*=.05). Multiple comparison *p*-values were corrected using the Benjamini-Hochberg procedure. Details on model development are reported in the supplementary materials.

## Results

### Cohort Characterization

Details regarding excluded cases and final cohort composition are reported in Figure S3 to Figure S10. Cohort characterization is available in Table S1.

### Report-Guided Corrections

After case exclusions, the unlabeled training data (PROMIS and PNET-INT-RETRO) comprised 7,340 (3,387 csPCa) cases. Using the FSL teacher model (trained on 2,700 cases), pseudolabels were retained for 2,483/3,387 (73.31%) csPCa cases. In contrast, the LOC teacher (trained on 1,150 positive cases) retained pseudolabels for 2,722/3,387 (80.37%) csPCa cases. Additional details are reported in Figure S11 and Figure S12.

### Lesion-Level Evaluation

Following exclusions, PI-CAI-VAL was left with 338 cases (338 patients), 105 of which had csPCa (111 connected components). Figure 2 depicts the fROC curves (left) and DSC values (right, [n=111]) for each of the used approaches. The fROC analysis demonstrates that, comparing to FSL, SSL methods reduce the number of false positives per case. This difference is more evident in the [0.6, 0.8] sensitivity interval. Regarding segmentation quality, RG-SSL-LOC yielded a median (IQR) DSC of 0.49 (0.36-0.61), significantly higher than the ones computed with FSL (0.41 [0.31-0.54], p<.001) and RG-SSL (0.40 [0.30-0.55], p<.001).

**Figure 2.**
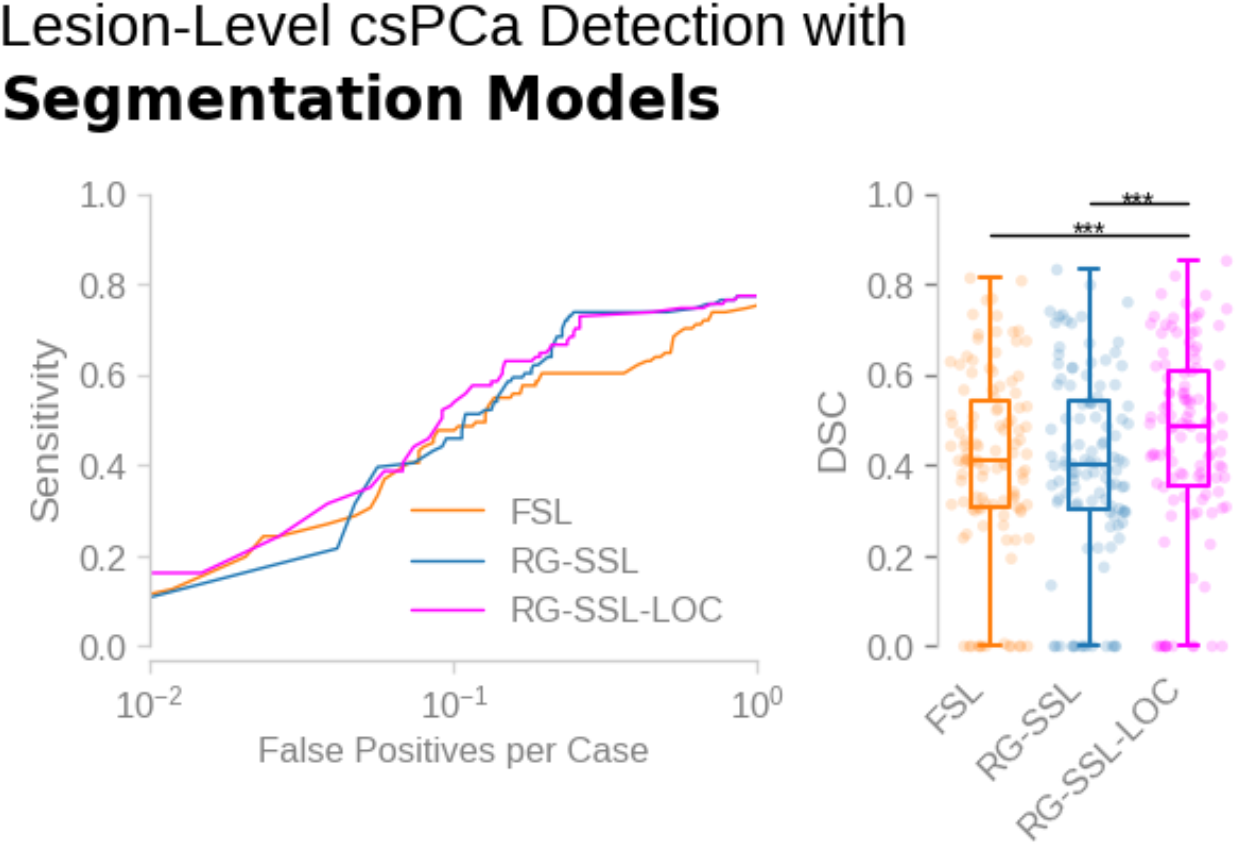
Lesion-level fROC analysis (left) and DSC values (right) computed on 111 connected components. *p < .05. **p <.01. ***p < .001.

### Case-Level Evaluation

#### A. Segmentation Models

The FSL, RG-SSL and RG-SSL-LOC csPCa segmentation models were trained on 2,700 (34.07% csPCa), 9,136 (37.35% csPCa) and 9,375 (38.94% csPCa) cases, respectively. Figure 3 (a) depicts the computed metrics for the External Retrospective (PNET-EXT-RETRO, n=515), External Prospective (PNET-EXT-PROS, n=590) and Internal Prospective (PNET-INT-PROS, n=1,336) cohorts.

**Figure 3.**
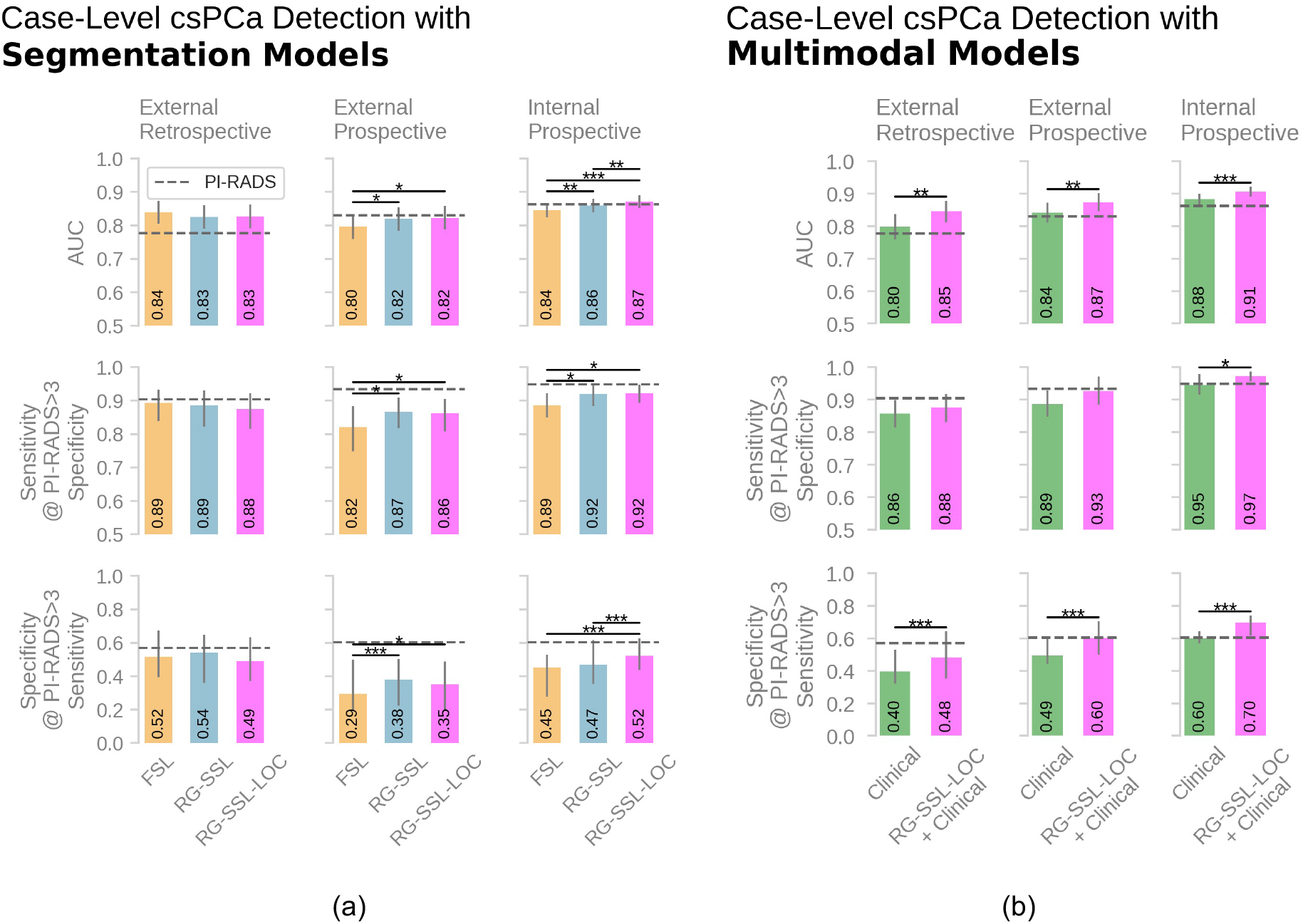
Case-level AUC, sensitivity and specificity values for csPCa detection with segmentation (a) and multimodal (b) models. PI-RADS metrics are depicted horizontal grey dashed lines. Error bars represent 95% confidence intervals. Statistically significant differences between AUCs were assessed using DeLong’s test, while differences in specificity values were evaluated using the Wilcoxon signed-rank test (both *alpha* = 0.05). *p < .05. **p <.01. ***p < .001.

In general, the SSL methods computed higher metrics than FSL, with the exception of the AUCs for the external retrospective cohort, where FSL achieved 0.84 (95% CI: 0.81-0.87), with no significant differences from RG-SSL (0.83 [95% CI: 0.79-0.86], p=.138) and RG-SSL-LOC (0.83 [95% CI: 0.79-0.86], p=.237). Although RG-SSL scored the highest sensitivity (0.89 [95% CI: 0.82-0.93]), it did not differ significantly from the other two methods, as well as PI-RADS (0.91 [95% CI: 0.87-0.94], p=.612). The same was observed for specificity, where RG-SSL scored 0.54 (95% CI: 0.36-0.65), not significantly different from PI-RADS (0.57 [95% CI: 0.51-0.63], p=.490).

Both SSL approaches achieved significantly higher performance metrics than FSL on the external prospective dataset, with no significant differences observed between the two. Compared with the external retrospective dataset, the reductions in AUC, sensitivity and specificity were smaller for RG-SSL (0.01, 0.02 and 0.16, respectively) and RG-SSL-LOC (0.01, 0.02 and 0.14, respectively) than for FSL (0.04, 0.07 and 0.23, respectively), suggesting improved robustness of the SSL approaches.

On the internal prospective set, RG-SSL-LOC yielded an AUC of 0.87 (95% CI: 0.85-0.89), significantly higher than both FSL (0.84 [95% CI: 0.82-0.87], p<.001) and RG-SSL (0.86 [95% CI: 0.84-0.88], p=.007), but not significantly higher than PI-RADS (0.86 [95% CI: 0.85-0.88], p=.480). Similarly, the scored specificity was also significantly higher than the other two methods (p<.001). While sensitivity did not differ significantly between RG-SSL and RG-SSL-LOC (0.92 [95% CI: 0.88-0.95] and 0.92 [95% CI: 0.89-0.95], respectively, p=.796), both were significantly higher than FSL (0.89 [95% CI: 0.85-0.92], both p=.010).

Subgroup analysis (Figure S14 to Figure S18) showed that, in most cases, that difference between retrospective and prospective AUCs was smaller for RG-SSL-LOC than for FSL. Based on the lesion- and case-level results for csPCa, RG-SSL was excluded from further analysis. Consequently, the FSL and RG-SSL-LOC PCa segmentation models were trained on 1,369 (60.99% PCa) and 7,723 (57.65% PCa) cases, respectively. The corresponding results are presented in Figure S19 (a). Overall, metrics were substantially lower than those achieved with PI-RADS. Nevertheless, RG-SSL-LOC consistently outperformed FSL across all validation cohorts, with statistically significant improvements in both external retrospective and internal prospective cohorts.

#### B. Multimodal Models

After excluding cases with missing PI-RADS, each csPCa detection ensemble was trained with 711 cases distributed over five LR models. The resulting metrics are depicted in Figure 3 (b). *Clinical* refers to the multimodal model trained on clinical variables alone, whereas *RG-SSL-LOC+Clinical* denotes the multimodal model trained on both clinical variables and *P*_*RG-SSL-LOC*_*(csPCa), i*.*e*. the case-level csPCa likelihood estimated with RG-SSL-LOC. *RG-SSL-LOC+Clinical* consistently scored significantly higher metrics across all sets when compared to *Clinical*. The AUC improvement over PI-RADS was significant across sets (p≤.002 for all cases). In contrast, *RG-SSL-LOC+Clinical* did not achieve significantly higher sensitivity than PI-RADS across cohorts, but scored significantly higher specificity in the internal prospective cohort (0.70 [95% CI: 0.62–0.74] vs. 0.61 [95% CI: 0.57–0.64], p<.001) translating into 15.19% fewer unnecessary biopsies (599 and 520 true negatives for *RG-SSL-LOC+Clinical* and PI-RADS, respectively). DCA (Figure S13) confirmed the advantage of RG-SSL-LOC over a PI-RADS>3 decision threshold in this cohort.

The mean LR coefficients are shown in Figure 4, where *P*_*RG-SSL-LOC*_*(csPCa)* exhibited the highest mean coefficient (1.10 [95% CI: 0.49-1.71]) and remained statistically significant across all models.

**Figure 4.**
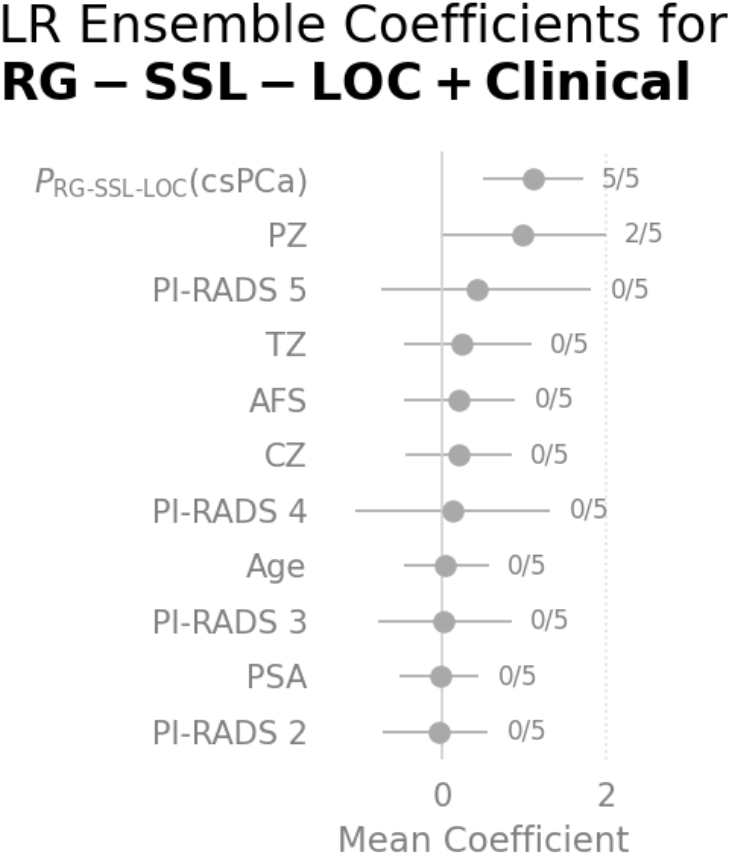
Mean LR ensemble coefficients (and corresponding mean 95% CIs) across fold models. Annotations on the right report the number of folds in which the corresponding p-value was < .05. P_RG-SSL-LOC_(csPCa) is the case-level csPCa likelihood estimated with the RG-SSL-LOC approach. AFS = anterior fibromuscular stroma, CZ = central zone, PZ = peripheral zone, TZ = transition zone. PI-RADS 1 was omitted due to redundancy following one-hot encoding.

The PCa ensembles were trained with 588 cases. The respective results are reported in Figure S19 (b). Although *RG-SSL-LOC+Clinical* scored higher metrics than *Clinical*, it fell short of the PI-RADS score, having only scored higher AUC for the external retrospective and internal prospective cohorts (p=.021 and p<.001, respectively).

## Discussion

Overall, the report-guided methodology increased the amount of training data by more than threefold, substantially reducing the annotation burden while facilitating scaling. The results are consistent with Bosma *et al*. (2025), with both SSL approaches achieving comparable improvements over FSL. Compared to the FSL teacher, our proposed LOC teacher retained additional 239 malignant cases, corresponding to a 7.02% increase in the total number of malignant cases available for training. Despite the modest increase, this translated into significantly improved segmentation performance.

At case-level, although both SSL csPCa segmentation models outperformed FSL on most validation cohorts, neither approach significantly surpassed the other on external data. The fact that the proposed method demonstrated an advantage only on the internal prospective cohort suggests that retaining a larger number of unlabeled malignant cases appears to primarily benefit detection on prospective cases from centers represented in the training set. The subgroup analysis demonstrated how, in general, RG-SSL-LOC is more robust to distribution shifts between retrospective and prospective external data, suffering from less AUC degradation in most cases across centers, scanner manufacturers (particularly relevant for General Electric scanners, which were overrepresented in the validation data relative to training), magnetic field strength, age and PSA.

Case-level AUCs achieved by multimodal csPCa models demonstrate the added value of incorporating segmentation-derived likelihood. At the PI-RADS>3 sensitivity operating point, RG-SSL-LOC improved specificity over PI-RADS only on the internal prospective cohort, potentially reducing unnecessary biopsies by 15.19%. Although this value differs from those reported in previous studies using different models — 20.2% (9) and 21.9% (10) —, those studies evaluated different validation cohorts, both in terms of participating centers and number of cases.

Although the SSL segmentation models were trained on >9,000 cases, the multimodal ensemble models were trained on only 711 cases (588 for PCa) to avoid data leakage (see supplementary materials). This comparatively less diverse training set may therefore have limited the generalizability of the multimodal models to external data.

Regarding PCa detection, although the proposed method outperformed FSL, it remained inferior to PI-RADS and achieved worse performance than its csPCa counterparts. This difference is likely attributable to the greater difficulty of detecting ISUP 1 lesions, which are typically smaller and less conspicuous on MRI.

This study is not devoid of limitations. The absence of internal retrospective cohort prevents a direct assessment of the impact of temporal distribution shifts within training centers. Besides, lesion segmentations were only available for retrospective validation data and lesion-level evaluation could not be performed prospectively. While we consider PI-RADS>3 as the biopsy referral threshold, PI-RADS=3 is equivocal and may warrant the assessment of other factors (PSA level, family history, patient preference), indicating that other PI-RADS thresholds should be tested in future research. While our zonal segmentation model performs well, it does not allow the segmentation of finer regions (*i*.*e*. CZ, TZ, AFS), leading to less precise pseudolabel allocation. Additionally, small errors in zonal probability maps might lead to downstream errors in lesion prediction. The lack of information regarding radiologist years of experience prevents assessing how the proposed methods compare with readers of different expertise levels. Furthermore, our dataset was collected from cases from clinical practice, having a single PI-RADS score for each case; this hindered the estimation of the inter-rater agreement, a more suitable metric of comparison for our models. Although prediction uncertainty is relevant to clinical implementation, its assessment was beyond the scope of this work. Nevertheless, our ensemble-based approach enables its quantification through metrics such as entropy.

In conclusion, we built upon the framework by Bosma *et al*. (2025) and proposed: 1) prospective multicenter validation; 2) multimodal integration; 3) csPCa vs. PCa comparison; 4) a lesion-only teacher model trained exclusively on ISUP≥1 cases to generate location-corrected pseudolabels. In general, RG-SSL-LOC approach made more effective use of unlabeled data, improving segmentation, enhancing robustness to prospective data and distribution shifts, and consistently improving multimodal detection.

Overall, SSL can effectively leverage large collections of MRI examinations, reducing reliance on manual annotations while supporting the development of scalable, robust, and clinically applicable AI systems. However, it is paramount to ensure that continued prospective validation and integration within appropriate MLOps frameworks allows their safe implementation in clinical practice (11).

## Data Availability

The PI-CAI and UCLA datasets are publicly available through their respective repositories. The PROMIS dataset is available upon reasonable request. The ProstateNET dataset was collected within the ProCAncer-I project and is not publicly available due to patient privacy, data protection regulations, and consortium data-sharing agreements.

https://zenodo.org/records/6624726

https://ncita.org.uk/promis-data-set-open-access-request/

