## Supplementary Materials for "Report-Guided Semi-Supervised Learning for Scalable Prostate Cancer Detection on Biparametric MRI: Multicenter Prospective Validation and Multimodal Integration"

### Data Splitting

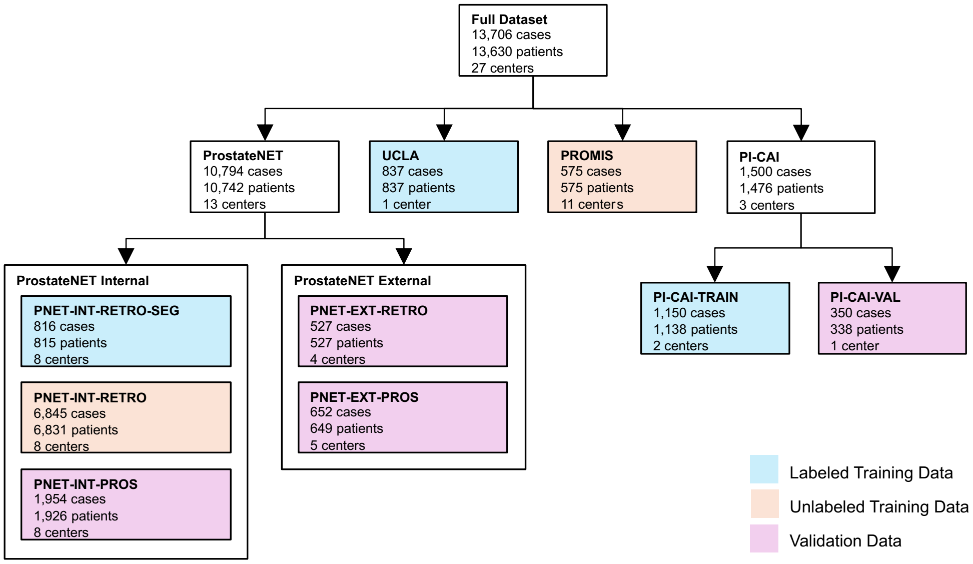


Figure S1: Overview of the division of the full dataset into training and validation cohorts. The reported numbers are before case exclusions. *Internal* and *external* refer to data from centers included in and excluded from the training sets, respectively.

### Data Preprocessing

All DWI and ADC sequences were resampled to T2w space. Peripheral and central/transitional zone segmentation masks were derived from the T2w sequences using an in-house model (1). Simple heuristic rules were applied to automatically exclude unreliable masks. Cases were excluded if: 1) a mask could not be generated for either zone; 2) the generated masks did not intersect the central *x*-coordinate of the image, allowing for a 10% margin on both the left and right sides; 3) if the connected components from the two zones did not remain in contact following a 3-dimensional binary dilation. To reduce the amount of non-prostatic signal ingested by the models, a 3-dimensional crop centered in the whole-gland mask (inferred from the zonal masks) was applied to all volumes and masks, keeping a 20% margin on both sides from each axis to account for periprostatic adipose tissue, which may play a role in PCa progression (2). To homogenize location priors, Barzell zones were converted to PI-RADS v2 sectors following the mapping proposed by Satish et al. (3). Since the UCLA dataset included biopsy coordinates, each associated with an ISUP grade, this information was used to filter the provided lesion segmentations according to the detection task. Specifically, each biopsy trajectory was modeled as a cylinder with a 1.2 mm diameter, corresponding to an approximate diameter (4) of the biopsy needle present in the system used by the authors (18-gauge (5)). The lesion segmentations were then matched to the biopsy trajectory cylinders based on the highest spatial overlap, thus linking each lesion to an ISUP grade.

For all validation cohorts, only the earliest MRI examination from each patient was retained. This prevented multiple examinations from the same patient from contributing to model evaluation. The earliest examination was selected to reflect the patient's initial clinical presentation.

### Model Development

We developed models for segmenting csPCa and PCa lesions present in bpMRI volumes. For each task, the predictions of the best performing segmentation model were then combined with clinical variables — PI-RADS score, age, PSA level, and lesion location — to train a clinical multimodal model for case-level detection. Both segmentation and multimodal models were trained and validated on a workstation equipped with 2x NVIDIA GeForce RTX 3090 (24GB VRAM) GPUs, Intel Xeon(R) W-2223 quad-core CPU and 256GB DDR4 RAM.

### Segmentation Models

All segmentation models were trained using the nnU-Net self-configuring framework (6). The input consisted of five channels: the preprocessed T2w, DWI and ADC volumes, together with the generated probability maps for the peripheral and central/transitional zones (based on the findings from Hosseinzadeh *et al*. (7)). All networks were implemented using the 3D full resolution architecture and trained for 1000 epochs (250 mini-batches per epoch). All nnU-Net default settings were kept, except for the loss function. Since initial experiments showed that the predicted probabilities were concentrated around the extremes (0 and 1), resulting in poor calibration, preventing meaningful threshold variation for ROC and fROC analysis. The standard loss function was replaced with the one proposed by Murugesan *et al*. (8), which includes label smoothing and margin loss. A smoothing factor $\alpha$ of 0.1 and a margin of 10 were selected empirically.

Training followed a 5-fold Cross-Validation (CV) strategy. For SSL models, only manually labeled cases were included in each validation fold, ensuring fold performance was evaluated against reliable ground-truth (Figure S2 [a]). During inference, each of the five trained models generated a probability map *P*, which was averaged voxel-wise to obtain the final ensemble prediction. Lesion candidates were then extracted from *P* using the same methodology as Bosma *et al*. (2023) (9). The first lesion candidate was defined as the voxel with highest probability, together with all neighboring voxels whose probability is at least 40% of this peak value. The corresponding region was set to zero and this procedure was repeated iteratively until a maximum of five lesion candidates were identified or no candidate remained. Each candidate was required to have a size of 10 or more voxels and have a maximum probability value higher than 0.01. The case-level likelihood of csPCa (or PCa) was defined as the maximum probability across lesion candidates.

### Multimodal Models

All clinical multimodal models — *i.e.*, models incorporating clinical variables — were based on Logistic Regression (LR) classifiers, implemented using *statsmodels* Python library (version 0.14.6). We used a maximum of 1000 iterations, L2 penalty and the Limited-memory BFGS (Broyden–Fletcher–Goldfarb–Shanno) optimizer. Due to the limited availability of unseen data for hyperparameter tuning, LR was selected for its interpretability and low dependence on hyperparameters.

LR multimodal models were trained on top of the segmentation models using an ensemble strategy to prevent data leakage (Figure S2 [b]). For each nnU-Net fold, the maximum lesion probability was extracted from each case included in the validation fold and used to train a LR model, together with PSA level, age, PI-RADS, and lesion location. The latter was defined as the prostate zone(s) affected by the lesion — peripheral zone (PZ), transition zone (TZ), central zone (CZ) or anterior fibromuscular stroma (AFS). Both PI-RADS and lesion location were one-hot encoded. All variables were *z*-standardized according to the training fold's mean and standard deviation. Missing PSA and age values were imputed on both training and validation folds using a *k*-nearest neighbors (with *k*=5) imputer fitted on the training fold. The PCa/csPCa case-level likelihood was then computed as the mean probability returned by the five LR models. Since PI-RADS scores were not available for UCLA and PI-CAI, all clinical models were trained exclusively on PNET-INT-RETRO-SEG cases without missing PI-RADS score.

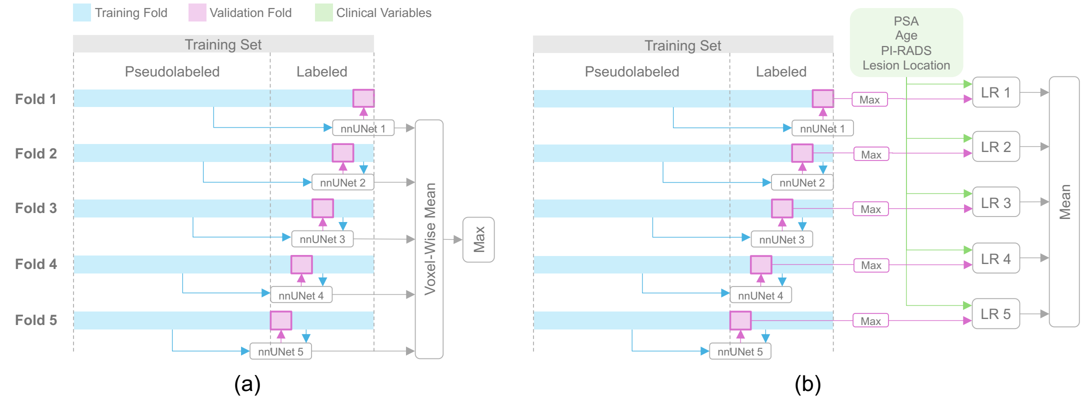


Figure S2: Training and inference strategies for the segmentation (a) and multimodal (b) models

### Cohort Characterization

Table S1: Cohort Characterization

| Variable | Training FSL | Training SSL | PNET-EXT-RETRO | PNET-EXT-PROS | PNET-INT-PROS | PI-CAI-VAL |
|---|---|---|---|---|---|---|
| **Age (Years)** | | | | | | |
| Median (IQR) | 65.0 (60.0-70.0) | 65.0 (60.0-70.0) | 66.0 (60.0-71.0) | 67.0 (61.0-72.0) | 65.5 (61.0-71.0) | 68.0 (63.0-71.8) |
| **PSA level (ng/mL)** | | | | | | |
| Median (IQR) | 7.3 (5.0-11.1) | 7.0 (5.0-10.6) | 6.4 (5.0-9.3) | 6.6 (5.0-9.6) | 6.7 (4.7-9.8) | 8.6 (6.2-12.0) |
| **PI-RADS Score** | | | | | | |
| 1 | 259 (9.6%) | 2133 (21.2%) | 84 (16.3%) | 75 (12.7%) | 458 (34.3%) | 0 (0.0%) |
| 2 | 11 (0.4%) | 85 (0.8%) | 8 (1.6%) | 61 (10.3%) | 28 (2.1%) | 0 (0.0%) |
| 3 | 314 (11.6%) | 687 (6.8%) | 72 (14.0%) | 88 (14.9%) | 58 (4.3%) | 0 (0.0%) |
| 4 | 478 (17.7%) | 2418 (24.1%) | 214 (41.6%) | 185 (31.4%) | 489 (36.6%) | 0 (0.0%) |
| 5 | 386 (14.3%) | 2164 (21.6%) | 137 (26.6%) | 181 (30.7%) | 303 (22.7%) | 0 (0.0%) |
| Missing | 1252 (46.4%) | 2553 (25.4%) | 0 (0.0%) | 0 (0.0%) | 0 (0.0%) | 338 (100.0%) |
| **csPCa** | | | | | | |
| Yes | 920 (34.1%) | 4307 (42.9%) | 273 (53.0%) | 246 (41.7%) | 478 (35.8%) | 105 (31.1%) |
| No | 1780 (65.9%) | 5733 (57.1%) | 242 (47.0%) | 344 (58.3%) | 858 (64.2%) | 233 (68.9%) |
| **Centre** | | | | | | |
| RADBOUDUMC | 947 (35.1%) | 3206 (31.9%) | 0 (0.0%) | 0 (0.0%) | 64 (4.8%) | 0 (0.0%) |
| HACETTEPE | 289 (10.7%) | 1957 (19.5%) | 0 (0.0%) | 0 (0.0%) | 393 (29.4%) | 0 (0.0%) |
| FCHAMPALIMAUD | 78 (2.9%) | 895 (8.9%) | 0 (0.0%) | 0 (0.0%) | 104 (7.8%) | 0 (0.0%) |
| NCI | 105 (3.9%) | 638 (6.4%) | 0 (0.0%) | 0 (0.0%) | 302 (22.6%) | 0 (0.0%) |
| RMH | 71 (2.6%) | 679 (6.8%) | 0 (0.0%) | 0 (0.0%) | 100 (7.5%) | 0 (0.0%) |
| UCLA | 738 (27.3%) | 738 (7.4%) | 0 (0.0%) | 0 (0.0%) | 0 (0.0%) | 0 (0.0%) |
| FPO | 56 (2.1%) | 549 (5.5%) | 0 (0.0%) | 0 (0.0%) | 184 (13.8%) | 0 (0.0%) |
| PROMIS | 0 (0.0%) | 528 (5.3%) | 0 (0.0%) | 0 (0.0%) | 0 (0.0%) | 0 (0.0%) |
| QUIRONSALUD | 37 (1.4%) | 325 (3.2%) | 0 (0.0%) | 0 (0.0%) | 87 (6.5%) | 0 (0.0%) |
| HULAFE | 0 (0.0%) | 0 (0.0%) | 187 (36.3%) | 211 (35.8%) | 0 (0.0%) | 0 (0.0%) |
| ZGT | 350 (13.0%) | 350 (3.5%) | 0 (0.0%) | 0 (0.0%) | 0 (0.0%) | 0 (0.0%) |
| PCNN | 0 (0.0%) | 0 (0.0%) | 0 (0.0%) | 0 (0.0%) | 0 (0.0%) | 338 (100.0%) |
| IDIBGI | 29 (1.1%) | 175 (1.7%) | 0 (0.0%) | 0 (0.0%) | 102 (7.6%) | 0 (0.0%) |
| UNIPI | 0 (0.0%) | 0 (0.0%) | 121 (23.5%) | 84 (14.2%) | 0 (0.0%) | 0 (0.0%) |
| GAONA | 0 (0.0%) | 0 (0.0%) | 0 (0.0%) | 185 (31.4%) | 0 (0.0%) | 0 (0.0%) |
| JCC | 0 (0.0%) | 0 (0.0%) | 107 (20.8%) | 55 (9.3%) | 0 (0.0%) | 0 (0.0%) |
| IPC | 0 (0.0%) | 0 (0.0%) | 100 (19.4%) | 55 (9.3%) | 0 (0.0%) | 0 (0.0%) |
| **Scanner Manufacturer** | | | | | | |
| Siemens | 2157 (79.9%) | 5708 (56.9%) | 137 (26.6%) | 88 (14.9%) | 183 (13.7%) | 69 (20.4%) |
| Philips | 343 (12.7%) | 2819 (28.1%) | 4 (0.8%) | 175 (29.7%) | 596 (44.6%) | 269 (79.6%) |
| General Electric | 165 (6.1%) | 1243 (12.4%) | 374 (72.6%) | 324 (54.9%) | 546 (40.9%) | 0 (0.0%) |
| Other | 1 (0.0%) | 8 (0.1%) | 0 (0.0%) | 1 (0.2%) | 0 (0.0%) | 0 (0.0%) |
| Missing | 34 (1.3%) | 262 (2.6%) | 0 (0.0%) | 2 (0.3%) | 11 (0.8%) | 0 (0.0%) |
| **Magnetic Field Strength (T)** | | | | | | |
| 1.5 | 455 (16.9%) | 3736 (37.2%) | 109 (21.2%) | 39 (6.6%) | 770 (57.6%) | 29 (8.6%) |
| 3.0 | 2211 (81.9%) | 6042 (60.2%) | 405 (78.6%) | 547 (92.7%) | 555 (41.5%) | 40 (11.8%) |
| Missing | 34 (1.3%) | 262 (2.6%) | 1 (0.2%) | 4 (0.7%) | 11 (0.8%) | 269 (79.6%) |
| **Total** | 2700 | 10040 | 515 | 590 | 1336 | 338 |

**Note:** Training FSL refers to the data used to train the fully-supervised csPCa segmentation model and Training SSL to the data used to train the semi-supervised approaches (before report-guided corrections). PSA = Prostate-Specific Antigen; PI-RADS = Prostate Imaging Reporting and Data System; csPCa = Clinically Significant Prostate Cancer; RADBOUDUMC = Radboud UMC; HACETTEPE = Hacettepe University, School of Medicine; FCHAMPALIMAUD = Champalimaud Foundation; NCI = National Cancer Institute; RMH = Royal Marsden Hospital; UCLA = UCLA Clark Urology Center; FPO = Candiolo Cancer Institute; QUIRONSALUD = Quirónsalud Hospital; HULAFE = La Fe Hospital; ZGT = Ziekenhuis Groep Twente; PCNN = Prostaat Centrum Noord-Nederland, IDIBGI = Institut D'Investigacio Biomedica De Girona; UNIPI = University of Pisa; GAONA = General Anti-Cancer and Oncological Hospital of Athens, JCC = JCC Diagnostic Imaging; IPC = Institut Paoli Calmettes

### STARD Diagrams

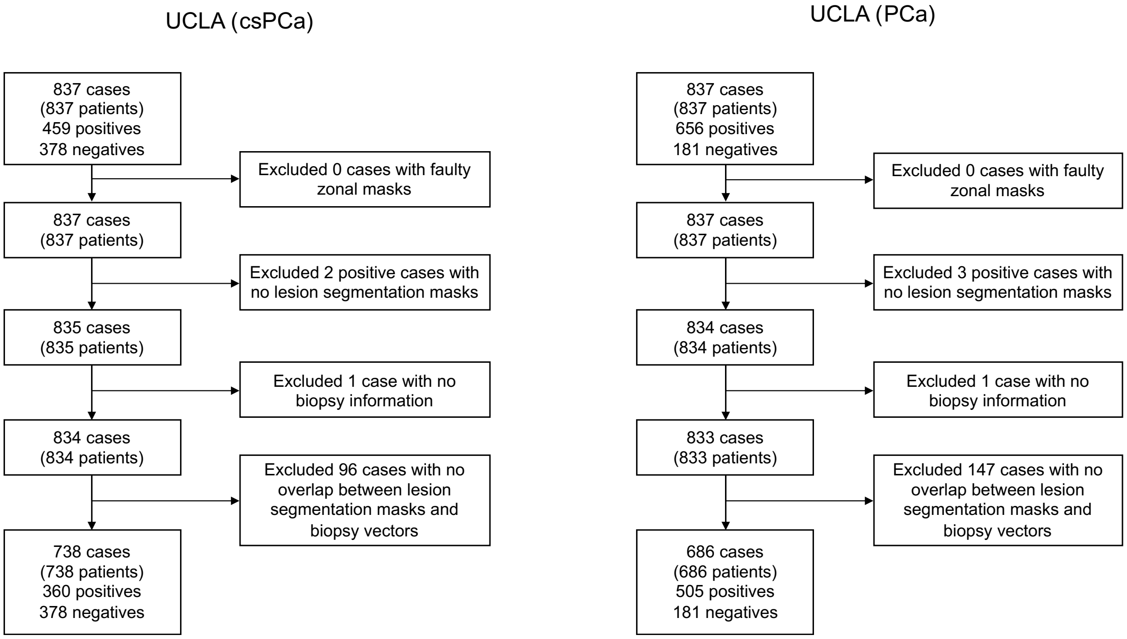


Figure S3: UCLA STARD Diagrams for csPCa (left) and PCa (right) detection tasks

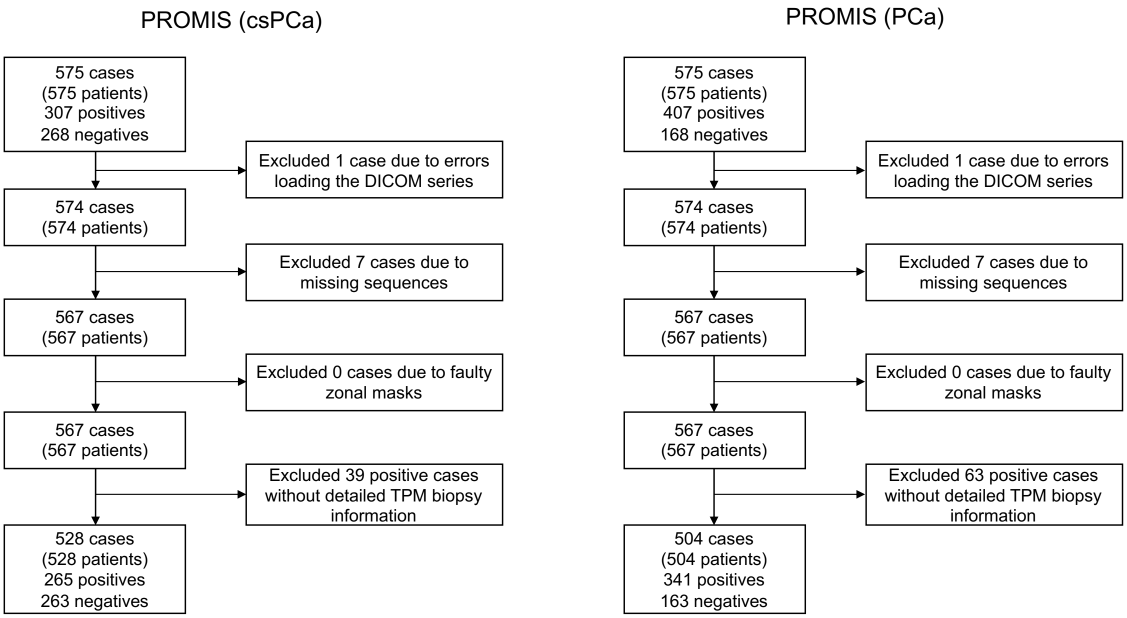


Figure S4: PROMIS STARD Diagrams for csPCa (left) and PCa (right) detection tasks

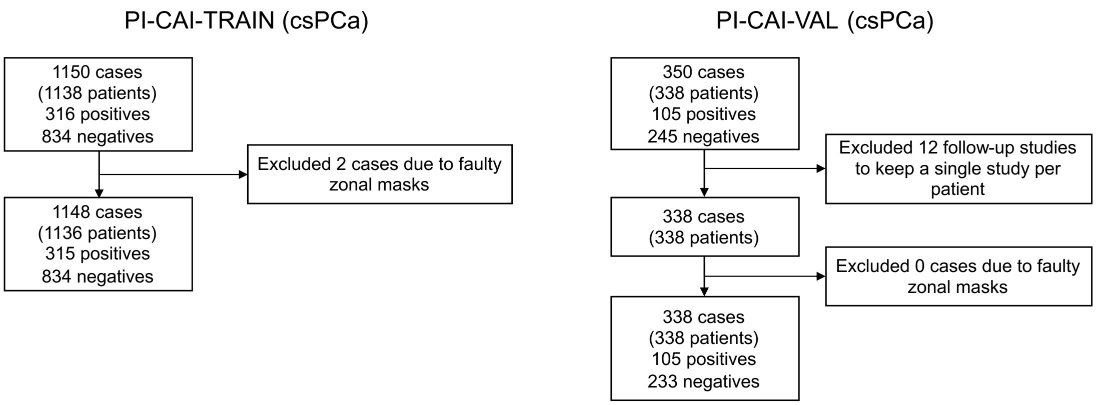


Figure S5: PI-CAI-TRAIN (left) and PI-CAI-VAL (right) STARD Diagrams for the csPCa detection task

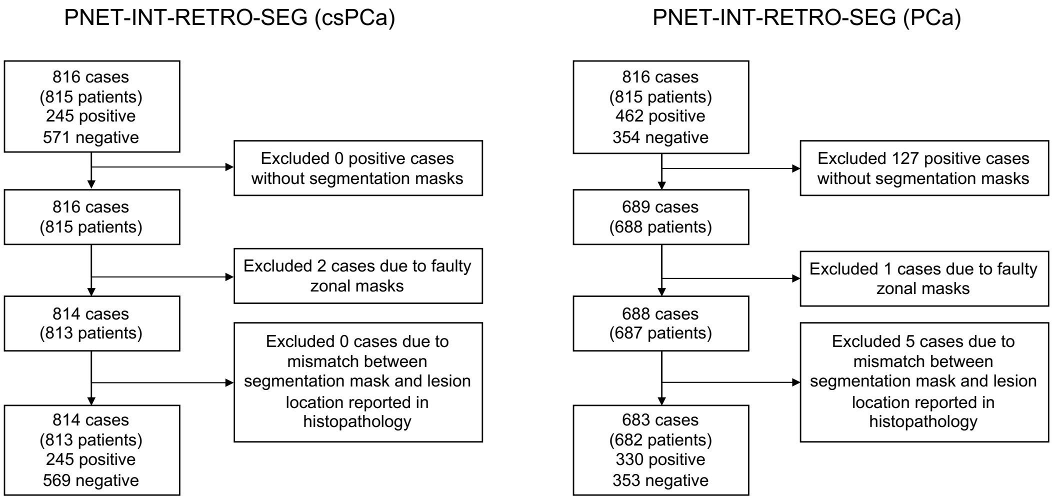


Figure S6: PNET-INT-RETRO-SEG STARD Diagrams for csPCa (left) and PCa (right) detection tasks

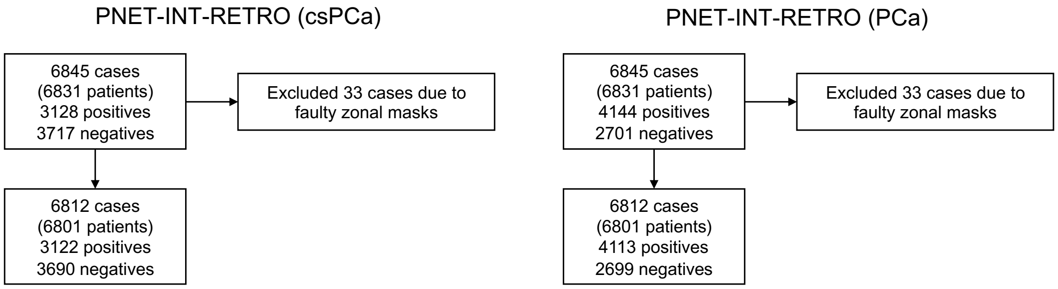


Figure S7: PNET-INT-RETRO STARD Diagrams for csPCa (left) and PCa (right) detection tasks

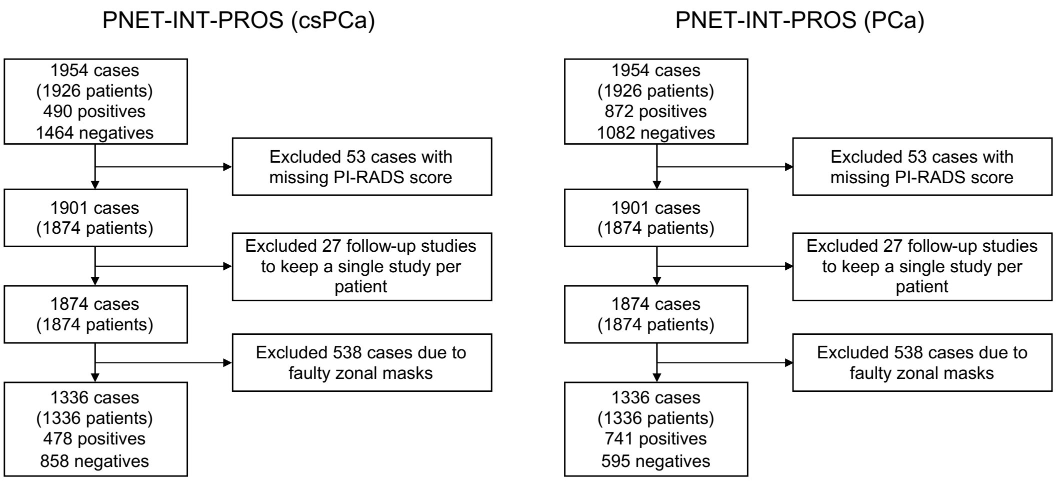


Figure S8: PNET-INT-PROS STARD Diagrams for csPCa (left) and PCa (right) detection tasks

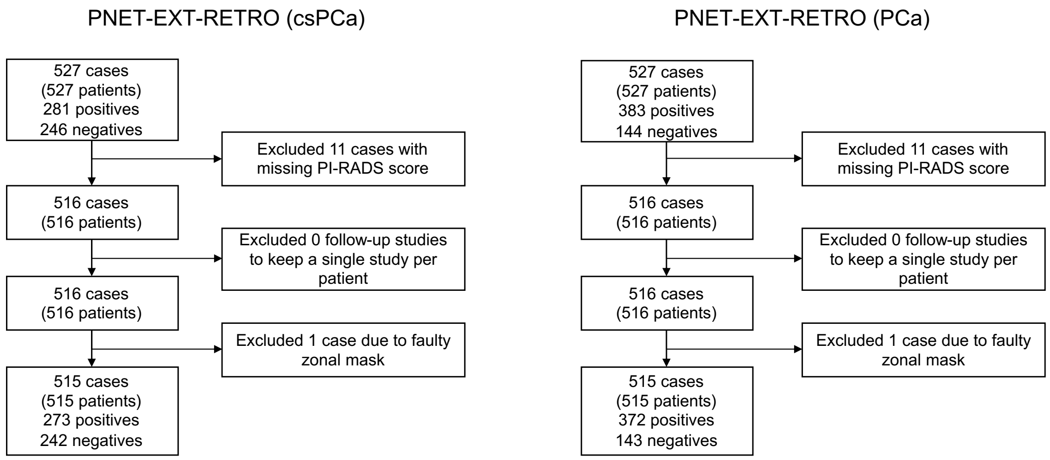


Figure S9: PNET-EXT-RETRO STARD Diagrams for csPCa (left) and PCa (right) detection tasks

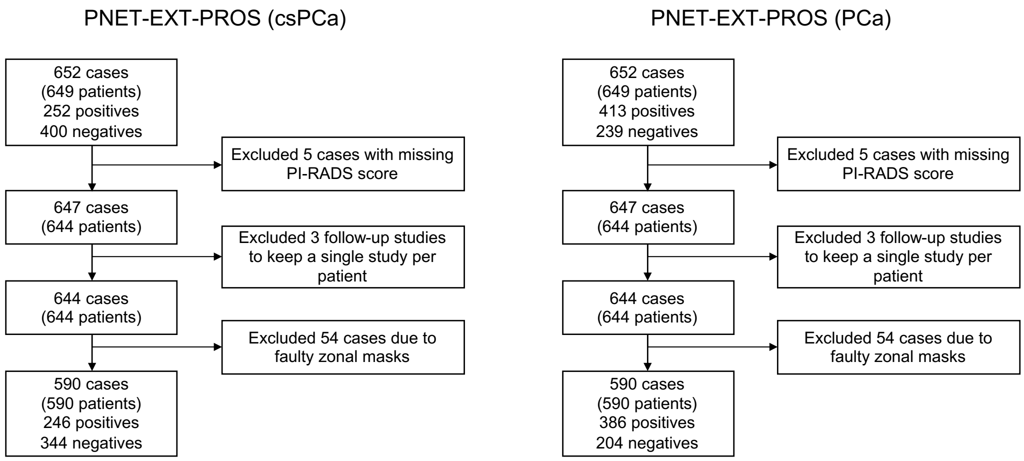


Figure S10: PNET-EXT-PROS STARD Diagrams for csPCa (left) and PCa (right) detection tasks

### Report-Guided Corrections

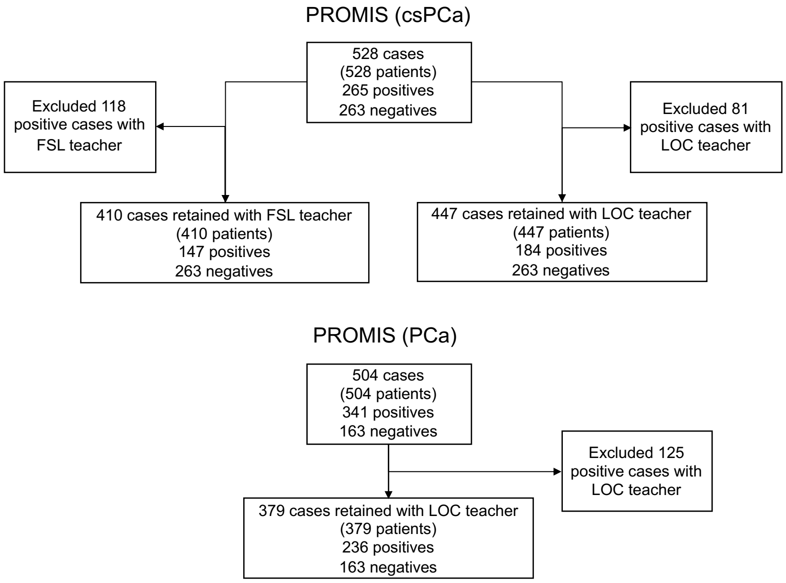


Figure S11: PROMIS report-guided corrections STARD diagrams for csPCa (top) and PCa (bottom) detection tasks

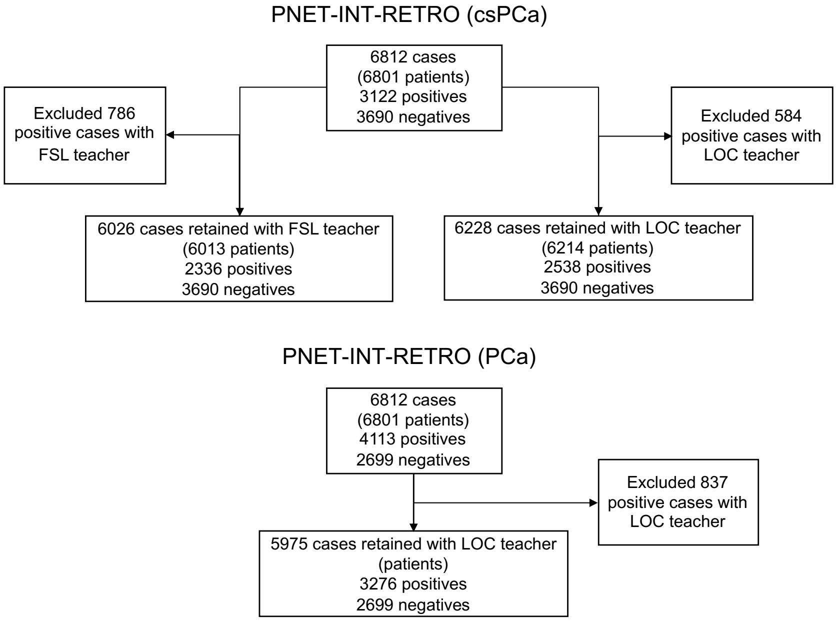


Figure S12: PNET-INT-RETRO report-guided corrections STARD diagrams for csPCa (top) and PCa (bottom) detection tasks

### Results on Case-Level csPCa Detection

Table S2: Mean AUC, sensitivity at the PI-RADS >3 specificity, and specificity at the PI-RADS >3 sensitivity calculated for all csPCa detection models. Reported P values represent comparisons between each model metric and the corresponding PI-RADS score. An asterisk (*) indicates statistical significance (P < .05).

| | **Model** | **External Retrospective** | **External Prospective** | **Internal Prospective** |
|---|---|---|---|---|
| **Mean AUC (95% CI)** | PI-RADS | 0.778 (0.740-0.816) | 0.831 (0.800-0.861) | 0.863 (0.846-0.880) |
| | FSL | 0.839 (0.805-0.873)<br>P=0.019* | 0.796 (0.759-0.833)<br>P=0.133 | 0.845 (0.823-0.866)<br>P=0.122 |
| | RG-SSL | 0.825 (0.789-0.861)<br>P=0.055 | 0.819 (0.784-0.854)<br>P=0.652 | 0.859 (0.839-0.880)<br>P=0.685 |
| | RG-SSL-LOC | 0.826 (0.790-0.862)<br>P=0.055 | 0.823 (0.788-0.858)<br>P=0.652 | 0.871 (0.852-0.890)<br>P=0.480 |
| | Clinical | 0.798 (0.760-0.837)<br>P=0.104 | 0.842 (0.811-0.873)<br>P=0.255 | 0.882 (0.865-0.900)<br>P<0.001* |
| | RG-SSL-LOC + Clinical | 0.845 (0.812-0.879)<br>P<0.001* | 0.874 (0.846-0.902)<br>P=0.002* | 0.906 (0.891-0.921)<br>P<0.001* |
| **Mean Sensitivity (95% CI)** | PI-RADS | 0.905 (0.869-0.939) | 0.935 (0.902-0.965) | 0.950 (0.930-0.968) |
| | FSL | 0.894 (0.840-0.932)<br>P=0.612 | 0.821 (0.748-0.884)<br>P<0.001* | 0.887 (0.849-0.922)<br>P<0.001* |
| | RG-SSL | 0.886 (0.822-0.930)<br>P=0.612 | 0.866 (0.819-0.909)<br>P=0.005* | 0.921 (0.884-0.947)<br>P=0.065 |
| | RG-SSL-LOC | 0.875 (0.815-0.922)<br>P=0.612 | 0.862 (0.809-0.905)<br>P=0.005* | 0.923 (0.893-0.947)<br>P=0.076 |
| | Clinical | 0.857 (0.814-0.899)<br>P=0.028* | 0.886 (0.848-0.929)<br>P=0.014* | 0.946 (0.916-0.979)<br>P=0.715 |
| | RG-SSL-LOC + Clinical | 0.875 (0.832-0.918)<br>P=0.236 | 0.927 (0.885-0.971)<br>P=0.670 | 0.973 (0.956-0.987)<br>P=0.072 |
| **Mean Specificity 95% CI)** | PI-RADS | 0.570 (0.507-0.632) | 0.605 (0.551-0.656) | 0.606 (0.573-0.639) |
| | FSL | 0.517 (0.395-0.672)<br>P=0.392 | 0.294 (0.192-0.500)<br>P<0.001* | 0.450 (0.277-0.529)<br>P<0.001* |
| | RG-SSL | 0.541 (0.359-0.648)<br>P=0.49 | 0.381 (0.224-0.502)<br>P<0.001* | 0.467 (0.355-0.617)<br>P<0.001* |
| | RG-SSL-LOC | 0.492 (0.372-0.633)<br>P=0.192 | 0.352 (0.195-0.488)<br>P<0.001* | 0.522 (0.437-0.629)<br>P<0.001* |
| | Clinical | 0.397 (0.321-0.529)<br>P<0.001* | 0.494 (0.441-0.599)<br>P<0.001* | 0.600 (0.569-0.642)<br>P=0.500 |
| | RG-SSL-LOC + Clinical | 0.483 (0.351-0.643)<br>P=0.018* | 0.602 (0.501-0.704)<br>P=0.914 | 0.698 (0.617-0.741)<br>P<0.001* |

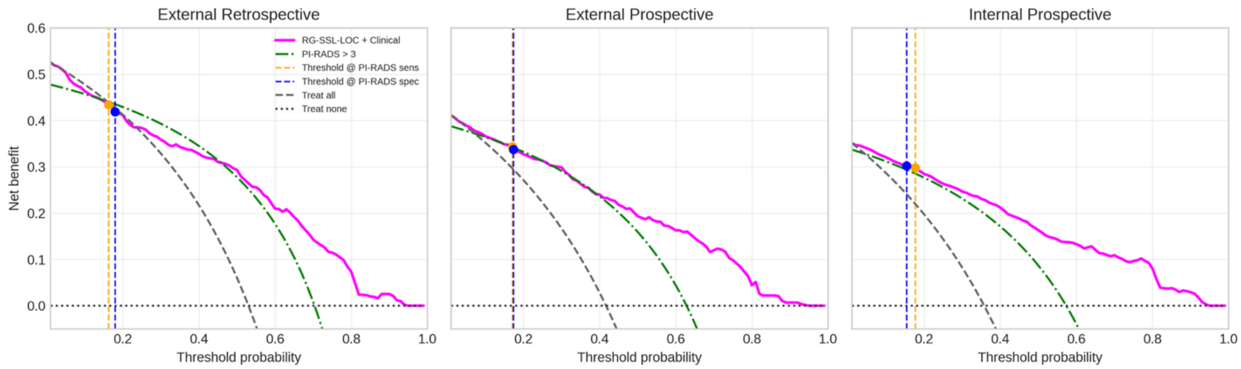


Figure S13: Decision curve analysis comparing RG-SSL-LOC+Clinical with PI-RADS < 3 for csPCa detection. The gray dashed and black dotted lines correspond to the “treat all” and “treat none” strategies, respectively. Orange and blue dashed vertical lines indicate the RG-SSL-LOC probability thresholds yielding the same sensitivity and specificity as PI-RADS > 3 specificity and sensitivity, respectively.

### Subgroup Analysis for Case-level csPCa detection

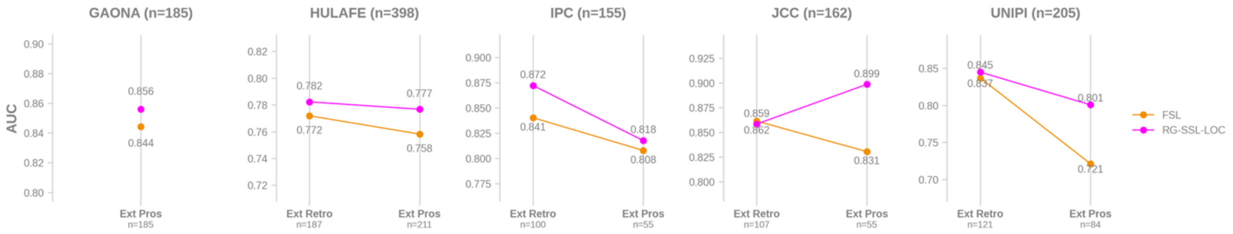


Figure S14: Subgroup analysis for centers (external retrospective and prospective)

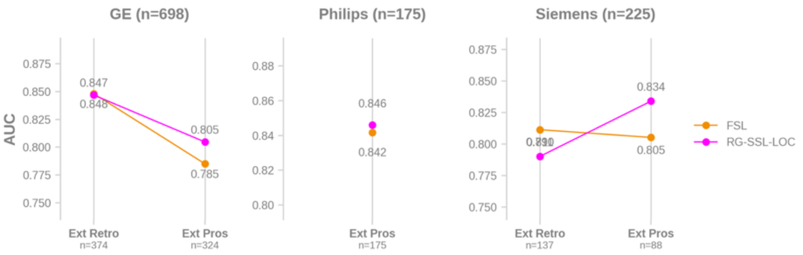


Figure S15: Subgroup analysis for scanner manufacturer (external retrospective and prospective)

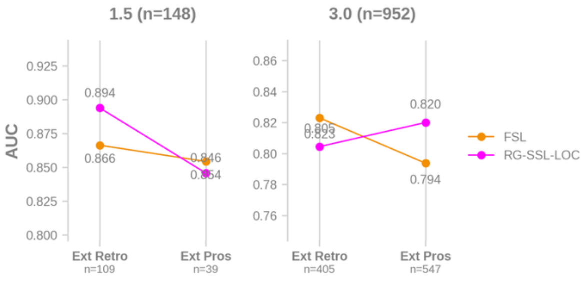


Figure S16: Subgroup analysis for scanner manufacturer (external retrospective and prospective)

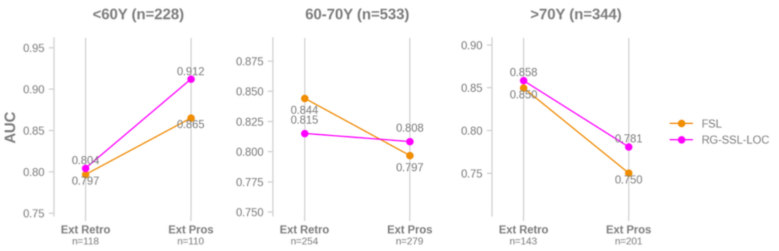


Figure S17: Subgroup analysis for age group (external retrospective and prospective)

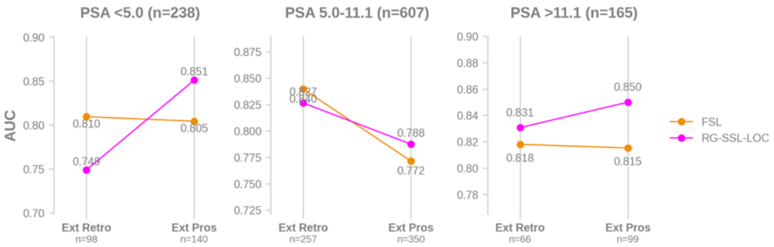


Figure S18: Subgroup analysis for PSA level group (external retrospective and prospective)

### Results on Case-Level PCa Detection

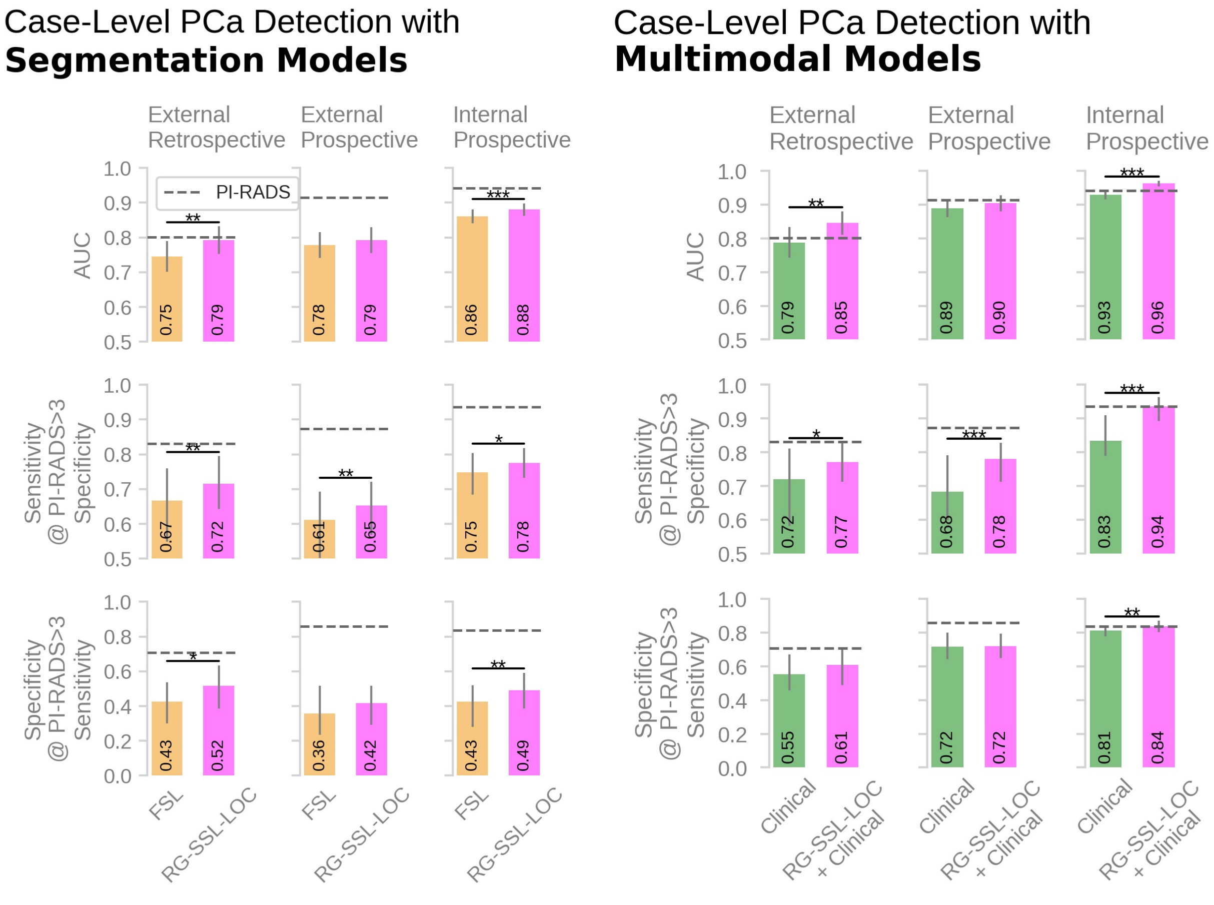


(a) (b)

Figure S19: Case-level AUC, sensitivity and specificity values for PCa detection with segmentation (a) and multimodal (b) models. PI-RADS metrics are depicted horizontal grey dashed lines. Error bars represent 95% confidence intervals. Statistically significant differences between AUCs were assessed using DeLong's test, while differences in specificity values were evaluated using the Wilcoxon signed-rank test (both alpha = 0.05). *p < .05. **p <.01. ***p < .001.

Table S3: Mean AUC, sensitivity at the PI-RADS >3 specificity, and specificity at the PI-RADS >3 sensitivity calculated for all PCa detection models. Reported P values represent comparisons between each model metric and the corresponding PI-RADS score. An asterisk (*) indicates statistical significance (P < .05).

| | Model | External Retrospective | External Prospective | Internal Prospective |
|---|---|---|---|---|
| Mean AUC (95% CI) | PI-RADS | 0.801 (0.758-0.843) | 0.914 (0.891-0.937) | 0.942 (0.930-0.953) |
| | FSL | 0.745 (0.702-0.789)<br>P=0.107 | 0.778 (0.740-0.816)<br>P<0.001* | 0.860 (0.840-0.880)<br>P<0.001* |
| | RG-SSL-LOC | 0.792 (0.752-0.832)<br>P=0.753 | 0.792 (0.755-0.829)<br>P<0.001* | 0.880 (0.862-0.898)<br>P<0.001* |
| | Clinical | 0.788 (0.743-0.833)<br>P=0.386 | 0.890 (0.863-0.917)<br>P=0.007* | 0.930 (0.916-0.944)<br>P=0.003* |
| | RG-SSL-LOC + Clinical | 0.846 (0.811-0.880)<br>P=0.021* | 0.904 (0.880-0.928)<br>P=0.298 | 0.962 (0.953-0.971)<br>P<0.001* |
| Mean Sensitivity (95% CI) | PI-RADS | 0.831 (0.792-0.867) | 0.873 (0.839-0.906) | 0.937 (0.919-0.953) |
| | FSL | 0.667 (0.549-0.760)<br>P<0.001* | 0.611 (0.503-0.693)<br>P<0.001* | 0.749 (0.685-0.804)<br>P<0.001* |
| | RG-SSL-LOC | 0.715 (0.643-0.795)<br>P<0.001* | 0.653 (0.569-0.721)<br>P<0.001* | 0.776 (0.733-0.819)<br>P<0.001* |
| | Clinical | 0.720 (0.582-0.812)<br>P<0.001* | 0.684 (0.599-0.791)<br>P<0.001* | 0.834 (0.790-0.909)<br>P<0.001* |
| | RG-SSL-LOC + Clinical | 0.772 (0.713-0.830)<br>P=0.006* | 0.780 (0.712-0.828)<br>P<0.001* | 0.935 (0.893-0.963)<br>P=0.903 |
| Mean Specificity 95% CI) | PI-RADS | 0.706 (0.630-0.779) | 0.858 (0.807-0.904) | 0.835 (0.806-0.865) |
| | FSL | 0.427 (0.300-0.536)<br>P<0.001* | 0.358 (0.233-0.518)<br>P<0.001* | 0.427 (0.280-0.520)<br>P<0.001* |
| | RG-SSL-LOC | 0.517 (0.385-0.634)<br>P=0.002* | 0.417 (0.291-0.518)<br>P<0.001* | 0.492 (0.387-0.591)<br>P<0.001* |
| | Clinical | 0.552 (0.456-0.671)<br>P=0.002* | 0.716 (0.643-0.799)<br>P<0.001* | 0.812 (0.778-0.843)<br>P=0.012* |
| | RG-SSL-LOC + Clinical | 0.608 (0.487-0.708)<br>P=0.065 | 0.721 (0.647-0.795)<br>P<0.001* | 0.835 (0.803-0.870)<br>P=1.000 |

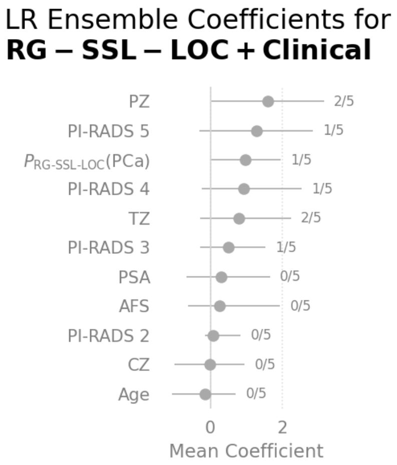


Figure S20: Mean LR ensemble coefficients (and corresponding mean 95% CIs) across fold models. Annotations on the right report the number of folds in which the corresponding p-value was < .05. $P_{\text{RG-SSL-LOC}}$(PCa) is the case-level PCa likelihood estimated with the RG-SSL-LOC approach. AFS = anterior fibromuscular stroma, CZ = central zone, PZ = peripheral zone, TZ = transition zone. PI-RADS 1 was omitted due to redundancy following one-hot encoding.
